# Causal relevance of different blood pressure traits on risk of cardiovascular diseases: GWAS and Mendelian randomisation in 100,000 Chinese adults

**DOI:** 10.1101/2023.01.20.23284709

**Authors:** Alfred Pozarickij, Wei Gan, Kuang Lin, Robert Clarke, Zammy Fairhurst-Hunter, Masaru Koido, Masahiro Kanai, Yukinori Okada, Yoichiro Kamatani, Yu Guo, Derrick Bennett, Huaidong Du, Yiping Chen, Ling Yang, Daniel Avery, Min Yu, Canqing Yu, Dan Schmidt Valle, Jun Lv, Junshi Chen, Richard Peto, Rory Collins, Liming Li, Zhengming Chen, Iona Y Millwood, Robin G Walters, the China Kadoorie Biobank (CKB) Collaborative Group

**Affiliations:** Clinical Trial Service Unit and Epidemiological Studies Unit (CTSU), Nuffield Department of Population Health, University of Oxford, Oxford, UK; Department of Computational biology and medical Sciences, Graduate School of Frontier Sciences, The University of Tokyo; Analytic and Translational Genetics Unit, Massachusetts General Hospital, Boston, MA, 02114, USA; Program in Medical and Population Genetics, Broad Institute of MIT and Harvard, Cambridge, MA, 02142, USA; Department of Statistical Genetics, Osaka University Graduate School of Medicine, Suita 565-0871, Japan; Department of Genome Informatics, Graduate School of Medicine, the University of Tokyo, Tokyo 113-0033, Japan; Laboratory for Systems Genetics, RIKEN Center for Integrative Medical Sciences, Kanagawa 230-150045, Japan; Laboratory of Statistical Immunology, Immunology Frontier Research Center (WPI-IFReC), Osaka University, Suita 565-0871, Japan; Medical Research Council Population Health Research Unit (MRC PHRU), Nuffield Department of Population Health, University of Oxford, Oxford, UK; National Center for Cardiovascular Diseases, Fuwai Hospital, Chinese Academy of Medical Sciences, Beijing, 100037,China; Zhejiang CDC, Zhejiang, China; Department of Epidemiology and Biostatistics, School of Public Health, Peking University, Beijing, China; China National Center For Food Safety Risk Assessment, Beijing, China; Peking University Center for Public Health and Epidemic Preparedness & Response

## Abstract

Elevated blood pressure (BP) is major risk factor for cardiovascular diseases (CVD). Genome-wide association studies (GWAS) conducted predominantly in populations of European ancestry have identified >2,000 BP-associated loci, but other ancestries have been less well-studied. We conducted GWAS of systolic, diastolic, pulse, and mean arterial BP in 100,453 Chinese adults. We identified 128 non-overlapping loci associated with one or more BP traits, harbouring 81 novel associations. Despite strong genetic correlations between populations, we identified appreciably higher heritability and larger variant effect sizes in Chinese compared with European or Japanese ancestry populations. Using instruments derived from these GWAS, multivariable Mendelian randomisation demonstrated strong causal associations of specific BP traits with CVD, including systolic BP with intracranial haemorrhage, and pulse pressure with carotid plaque. The findings reinforce the need for studies in diverse populations to understand the genetic determinants of BP traits and their role in disease risk.

## Introduction

Elevated blood pressure (BP) is a major modifiable cause of cardiovascular disease (CVD), including ischaemic heart disease (IHD) and stroke.^1-3^ In China, >4 million people develop CVD each year^4^ and the disease burden has increased steadily in recent decades, partially reflecting the rising prevalence of hypertension and suboptimal treatment of individuals with high BP.^5,6^ Randomised trials of several BP lowering medications have demonstrated proven efficacy for primary and secondary prevention of CVD.^7,8,9^ Although these BP lowering medications are widely used for primary and secondary prevention of CVD, their effectiveness varies for different CVD subtypes and according to overall proportions of stroke versus IHD.^10,11^

The most widely studied measures of BP are systolic BP (SBP) and diastolic BP (DBP), which reflect the force that the heart exerts on the arterial wall during the contraction and relaxation phases of the cardiac cycle, respectively. However, other measures of BP, including pulse pressure (PP) which reflects vascular stiffness, and mean arterial pressure (MAP) which reflects the average levels of blood pressure in arteries throughout the cardiac cycle, can provide additional insights into the determinants and consequences of BP.

For both individuals and populations, BP is determined by a complex interaction between environmental and genetic factors. Observational studies and randomised trials have demonstrated that lifestyle (e.g. alcohol,^12^ smoking,^13^ diet^14-19^), and environmental (e.g. ambient temperature^20^) factors influence levels of BP. Large genome-wide association studies (GWAS) have identified >2,000 genetic variants associated with different BP components, reflecting the high heritability of BP.^21-27^ However, most of the previous GWAS have been largely restricted to individuals of European ancestry, with limited data from Chinese and other East Asian ancestry populations, in which the genetic architecture, environmental exposures, and healthcare provision for treatment of high BP may differ substantially from those in Western populations. Moreover, most of the previous genetic studies have focused on SBP and DBP and, hence, relatively little is known about the genetic determinants of other BP traits including PP and MAP and their causal relevance for CVD.

In the present report, we examined GWAS findings for four different measures of BP in ∼100K participants in the China Kadoorie Biobank. Using summary statistics from European and Japanese cohorts, we compared BP heritability, genetic correlations, and SNP effect sizes on BP traits between Chinese, Japanese and European adults. In addition, using Mendelian randomisation (MR) approaches, we examined the causal relevance of different BP traits with risks of different vascular disease types in Chinese adults. We show that the causal relevance of BP traits varies for different CVD outcomes, potentially informing future investigation of the underlying biological mechanisms and development of anti-hypertensive therapies.

## Results

### Population characteristics

Of the 100,453 participants included in the GWAS (see **Supplementary Figure 1** for exclusion criteria), the mean age (SD) was 53.7 (11) years, mean BMI was 23.7 (3.5) kg/m^2^, 57.2% were women and 4.2% reported a prior history of CVD (**Supplementary Table 1**). Overall, men had a higher prevalence of hypertension (i.e. SBP ≥140 mmHg or DBP ≥90 mmHg at baseline recruitment, or self-reported prior diagnosis by a physician) than women (41.5% vs 36.6%). Among those with self-reported hypertension, approximately one-third reported the use of blood-pressure-lowering medication. Depending on the BP trait, age-specific mean BP measures in CKB were 0.5-7.7 mmHg lower than in populations of European ancestry (ICBP)^22^, but were comparable to those in Japanese adults (BBJ)^21^ (**Supplementary Table 2**). There were strong phenotypic correlations for BP phenotypes in CKB (**Supplementary Table 3**), even for pairs of traits that might be thought of as orthogonal (DBP and PP, PP and MAP).

### Genome-wide association analyses

We conducted GWAS of four BP traits (SBP, DBP, PP, MAP) both with and without adjustment for BMI (i.e. 8 separate GWAS). Although our primary analyses included adjustment for BMI, we conducted GWAS both with and without adjustment for BMI to facilitate comparisons with previous GWAS of these traits which have, variously, included or omitted adjustment for BMI. We identified a total of 128 non-overlapping genomic regions representing 510 separate trait-locus associations at *P*<5×10^−8^ (**Supplementary Tables 4-12**; **Supplementary Figures 2-5)**. Approximate conditional analyses identified 18 additional independent associations within 6 loci (**Supplementary Table 13**). Although there were minor differences between the BMI-adjusted and unadjusted analyses in the loci identified (120 and 115 loci, respectively, with 90 being identified in both sets of analyses), these almost entirely reflected small changes in statistical significance at or near the genome-wide significance threshold of 5×10^−8^. There were only very small differences between the two models in the estimated effect sizes for all lead variants, with the exception of a signal at the *FTO* locus, which showed a significantly reduced effect size in BMI-adjusted analysis for SBP, PP and MAP (**Supplementary Figure 6**).

Across the eight GWAS, there were 40 non-overlapping genomic regions corresponding to a total of 81 trait-specific associations which had not previously been reported in previous studies of BP in Europeans (ICBP^22,27^) or Japanese (BBJ^21,23,28,29^), or reported in the HGRI-EBI GWAS catalog^30^. A larger number of novel associations were identified for less well-studied MAP (24 and 21 with and without BMI adjustment, respectively) than for SBP (3 and 6), DBP (7 and 9), and PP (4 and 7) (**Supplementary Table 4**). One-third of the novel trait-locus associations were at loci previously reported as associated with at least one of the other BP phenotypes; after exclusion of these, 13 non-overlapping genomic regions identified as novel loci for a BP trait, comprising novel associations with one or more of SBP (2 and 4 with and without BMI adjustment, respectively), DBP (3 and 4), PP (2 and 3) and MAP (2 and 6). A larger proportion of genomic regions were newly-associated with one or more BP traits in East Asians, including 64 loci which had not previously been associated with any BP phenotype in East Asian populations (**Supplementary Table 4**).

### Comparisons between BP traits

Many of the identified loci were associated with multiple traits (**Figure 1a; Supplementary Figure 7**). Out of 120 BP-associated loci from the BMI-adjusted analyses, 22 (18%) were associated with all four BP traits, 29 (24%) with three BP traits, 21 (17%) with two BP traits, and 48 (40%) with only one BP trait. The extent of overlap was similar for associations from the BMI-unadjusted analyses. By contrast, of the 32 novel loci for BMI-adjusted traits, most of which were identified as associated with MAP, 26 (81%) were associated with one BP trait with the remaining 6 being associated with two traits (**Supplementary Figure 8**).

**Figure 1.**
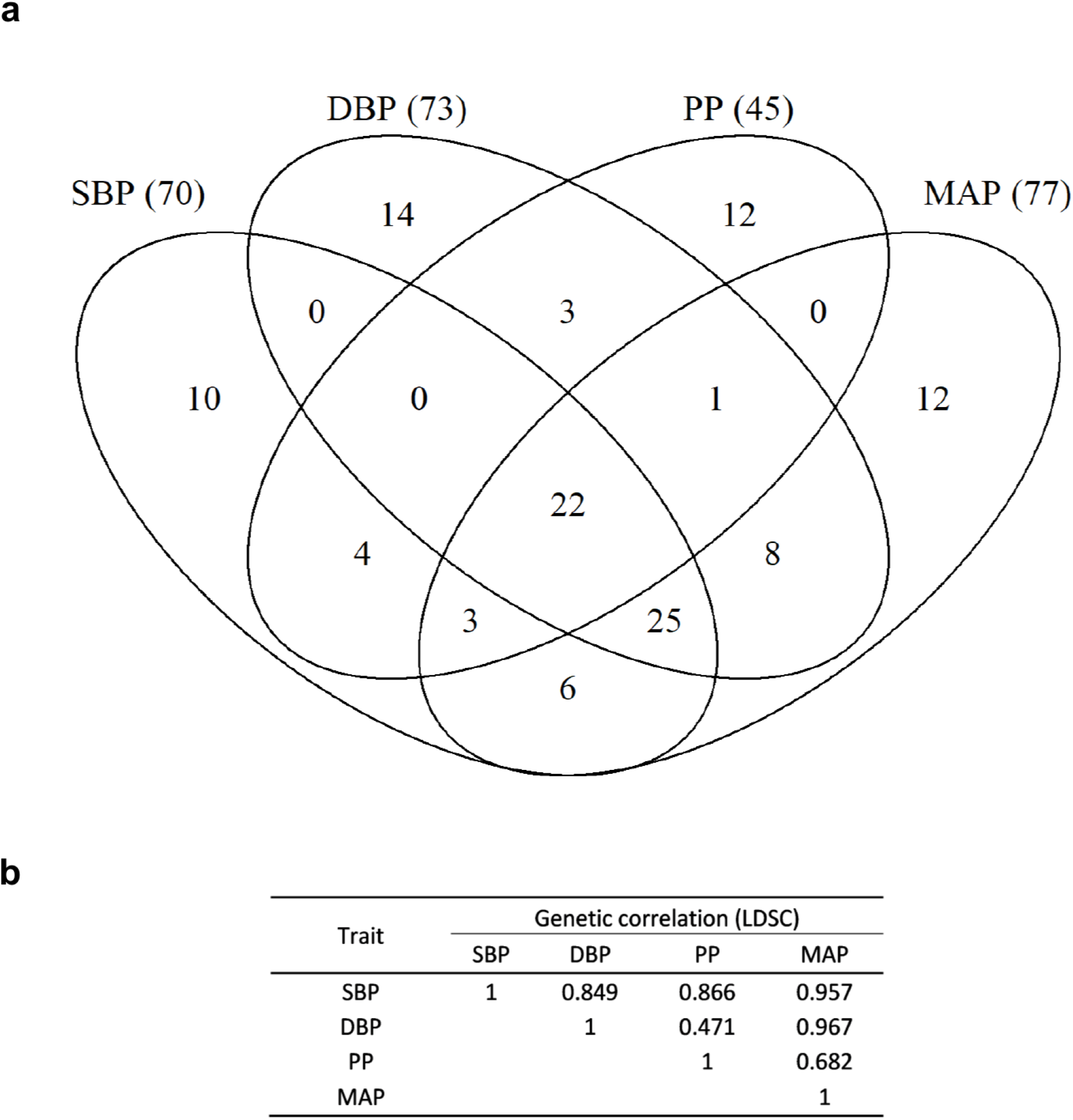
Overlap of associations across BMI-adjusted blood pressure traits. (a) Venn diagram of associations across BMI-adjusted blood pressure traits. The numbers in brackets indicate the total number of loci associated with that BP phenotype. (b) Genetic correlation between 4 traits as assessed by LD score regression.

The observational associations between BP traits (**Supplementary Table 3**) were reflected in the between-trait genetic correlations (**Figure 1b**). SBP and DBP showed strong genetic correlation with each other and with MAP (which is derived from them), with SBP also strongly correlated with PP. By comparison, reflecting its derivation as the difference between SBP and DBP, PP displayed substantially weaker genetic correlation with DBP (*r*_g_=0.471 [se 0.049]; BMI-unadjusted: *r*_g_=0.513 [0.039]) and with MAP (*r*_g_=0.682 [0.033]; BMI-unadjusted: *r*_g_=0.708 [0.026]). This is also reflected in comparisons of effect sizes for variants associated with each trait (**Supplementary Figure 9**): variants identified in GWAS of SBP, DBP, and MAP had effect sizes that were consistently correlated between traits, whereas variants identified in PP analyses showed very little correlation with the other traits. It was notable that variants consistently displayed larger effects on SBP than on the other traits, with regression lines having slopes of 1.76 [95%CI 1.72-1.80], 1.81 [1.73-1.90], and 1.44 [1.42-1.45], for comparisons with DBP, PP, and MAP, respectively. Similarly, variants had effect sizes for MAP (which is derived from SBP) which were consistently somewhat larger than for DBP and PP (1.25 [1.24-1.26] and 1.22 [1.12-1.29], respectively).

### Comparisons between populations

We sought to replicate the 81 novel variant-trait associations within 40 loci by performing association analyses in BBJ, and by performing lookups in published ICBP summary statistics^22^ (**Supplementary Table 14)**. A total of 77 associations were directionally consistent in BBJ, with association at 35 (43%) being replicated at 5% false discovery rate (FDR); similarly, 17 out of 23 available lead variants were directionally consistent in ICBP, with 9 replicated at 5% FDR. The replication analyses in BBJ consistently found variant effect sizes approximately one-third of those in CKB (**Supplementary Figure 10**). To investigate further, while avoiding the potential influence of “winners’ curse”^31^, we compared effect sizes in CKB with those published for BBJ, for lead variants identified for each trait in ICBP analyses (**Figure 2**). We again observed effect sizes overall to be substantially (1.7- to 2.1-fold) larger in CKB than for the corresponding associations in BBJ. The corresponding analyses comparing CKB and ICBP for lead variants identified in BBJ similarly showed 1.6- to 2.9-fold larger effect sizes in CKB. Consistent effect size differences were also found when separately comparing effect sizes in CKB participants recruited in urban (1.3- to 3.0-fold larger effect sizes in CKB) or rural (1.6- to 2.6-fold larger effect sizes in CKB) regions (**Supplementary Figure 11**).

**Figure 2.**
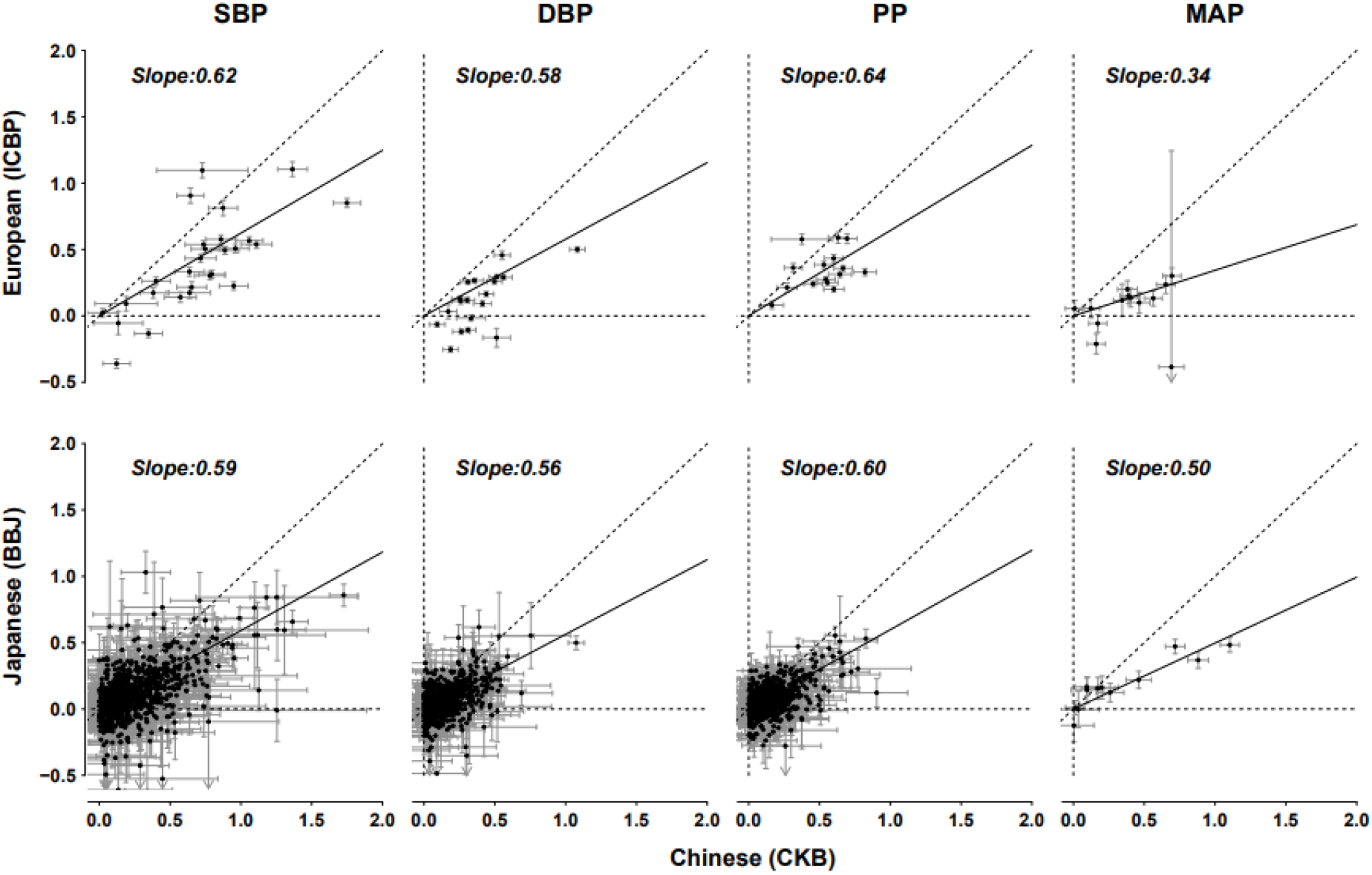
Comparisons of variant effect sizes of blood pressure traits between CKB and ICBP (BMI-adjusted) and between CKB and BBJ (BMI-unadjusted). Comparisons of CKB with ICBP and BBJ used variants identified in BBJ and ICBP, respectively. Variant per-allele effects are shown on the mmHg scale. The dashed diagonal line is the identity line (y = x). The solid line is the Deming regression line forced through the origin.

Using summary statistics from each of CKB, BBJ and ICBP, estimates of narrow-sense heritability (*h*^2^) using LD score regression were substantially and consistently higher in CKB compared with those observed for BBJ and ICBP, for all BP traits (**Table 1**). In particular, *h*^2^ for all BP traits was over 2-fold greater in CKB than in BBJ. There were also modest but consistent *h*^2^ differences between rural and urban CKB regions, with higher heritability in urban than rural regions (**Supplementary Figure 12, Supplementary Table 15**): for each BP phenotype, *h*^2^ in urban regions was 1.1- to 1.4-fold greater than in rural regions.

**Table 1.** Genome-wide significant associations and heritabilities for blood pressure traits in CKB.

| Trait | BMI adjustment | Loci | Novel associations | Heritability ($h^2_g$) | | |
| --- | --- | --- | --- | --- | --- | --- |
|  |  |  |  | CKB | BBJ | ICBP |
| SBP | - | 66 | 6 | 0.162 (0.010) | 0.076 (0.007) | - |
|  | + | 70 | 3 | 0.158 (0.012) | - | 0.129 (0.005) |
|  |  | **80** | **6** |  |  |  |
| DBP | - | 65 | 9 | 0.164 (0.010) | 0.060 (0.006) | - |
|  | + | 73 | 7 | 0.162 (0.011) | - | 0.129 (0.005) |
|  |  | **82** | **11** |  |  |  |
| PP | - | 45 | 7 | 0.129 (0.009) | 0.051 (0.005) | - |
|  | + | 45 | 4 | 0.124 (0.009) | - | 0.111 (0.004) |
|  |  | **51** | **8** |  |  |  |
| MAP | - | 69 | 21 | 0.171 (0.010) | 0.072 (0.007) | - |
|  | + | 77 | 24 | 0.168 (0.012) | - | 0.106 (0.009) |
|  |  | **86** | **31** |  |  |  |
| **Non-overlapping** |  | **128** | **40** |  |  |  |

SBP: systolic blood pressure; DBP: diastolic blood pressure; PP: pulse pressure; MAP: mean arterial pressure; CKB: China Kadoorie Biobank, BBJ: Biobank Japan; ICBP: International Consortium of Blood Pressure. Analyses were adjusted for sex, age, age<sup>2</sup>, research region, mean monthly external temperature and BMI adjustment. SNP-based heritability ( $h^2_g$ ) was estimated using LD score regression. Numbers in brackets are standard errors. Numbers in bold represent separate associations.

Despite these differences in variant effect size, for all BP traits (unadjusted for BMI) there was a high degree of between-population genetic correlation between CKB and BBJ (*r*_g_>0.870) as measured using both LD score regression and *Popcorn*^32^ (**Supplementary Table 16, Supplementary Table 17**). Similarly, there was strong cross-ancestry genetic correlation between CKB and ICBP for all traits (BMI-adjusted, *r*_g_>0.790) (**Supplementary Table 17**). By comparison, genetic correlations between BBJ (BMI unadjusted) and ICBP (BMI adjusted) were consistently somewhat lower (*r*_g_<0.75) for all traits.

### Associations of genetically-predicted blood pressure with major vascular diseases

Using independent lead variants for each trait, we constructed weighted genetic scores (GSs) for each (BMI-adjusted) BP trait for use in MR analyses of causal relationships between BP traits and major CVD outcomes (**Supplementary Table 18**). These explained 3.2%, 3.3%, 2.1%, and 3.6% of variance for SBP, DBP, PP, and MAP, respectively, with a tendency for somewhat larger r^2^ in rural regions. All instruments had an F-statistic of >100 for each region, indicating a very low risk of weak instrument bias. For all four traits, genetically-predicted BP showed strong positive associations with ischaemic stroke (IS), intracerebral haemorrhage (ICH), major coronary events (MCE) and carotid plaque (CP) (**Figure 3**). For IS, the odds ratio (OR) per 1-SD higher SBP (156 mmHg) was 1.84 [95% CI 1.61-2.10] higher risk, with other BP traits showing similar per 1-SD higher ORs. However, when effect sizes were instead expressed per 5 mmHg higher values for each trait, SBP showed the smallest (1.15, 1.11-1.18), while DBP showed the strongest (1.28, 1.21-1.35) association with IS. For ICH, the effects of genetically-determined BP were stronger than for IS, with SBP showing the strongest association per 1-SD higher BP (2.99, 2.50-3.57) but once again the smallest per 5 mmHg effect, for which DBP again showed the strongest association with ICH (1.51, 1.41-1.62). For MCE, the risk estimates were more modest, ranging from 1.34 to 1.38 per 1-SD higher BP. All four BP traits were significantly associated with CP, with genetically-predicted PP showing the strongest associations whether expressed per 1-SD (3.38, 2.57-4.46) or 5 mmHg higher (1.46, 1.34-1.59).

**Figure 3.**
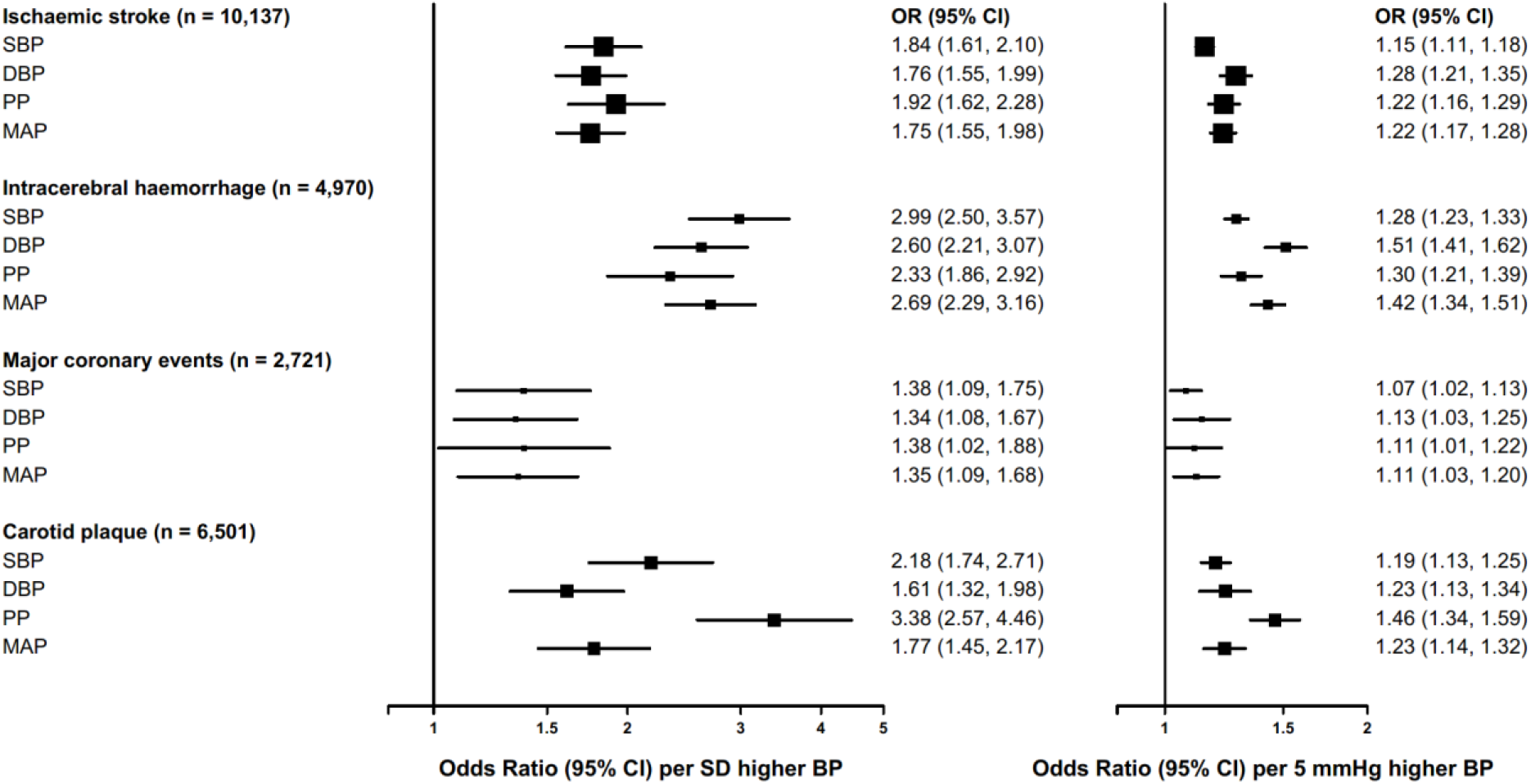
Mendelian randomisation of blood pressure traits with risk of major cardiovascular diseases and subclinical atherosclerosis. Effects are shown as odds ratios (95% CI) of disease risk per 1 SD higher blood pressure (left panel) and 5 mmHg higher blood pressure (right panel). SBP indicates systolic blood pressure; DBP, diastolic blood pressure; PP, pulse pressure; MAP, mean arterial pressure.

To distinguish true causal associations from those arising out of the strong genetic correlation and/or collinearity between the traits, we performed MVMR, in which pairs of instruments for different BP traits were included in a single model (**Figure 4, Supplementary Figure 13, Supplementary Table 19**). Each of these analyses used slightly different pairs of GSs, comprising variants at all loci that were associated with either of the pair of traits under investigation and, therefore, these GSs included a larger number of SNPs and explained a higher proportion of trait variance (2.4%-3.8%) than the instruments constructed for univariate MR. In the MVMR analyses, the different GSs for each trait gave univariate MR estimates which were consistent with each other and with the preceding trait-specific MR (**Figure 3**); however, when used in combination, so that they were mutually adjusted for each other, changes in effect estimates were observed which are interpreted as reflecting the causal relevance of individual BP traits for disease outcomes independent of contributions due to the other BP trait included in the analysis. For most pairs of traits, the conditional F-statistic was consistently greater than 10 for every recruitment region (F_min_>17), indicating a low risk of weak instrument bias; however, when MAP was analysed with either SBP or DBP, conditional F-statistics were often lower than 10, so that caution is required when interpreting the results of analyses using these pairings. The collinearity between these traits was reflected in wide confidence intervals in MVMR analyses.

**Figure 4.**
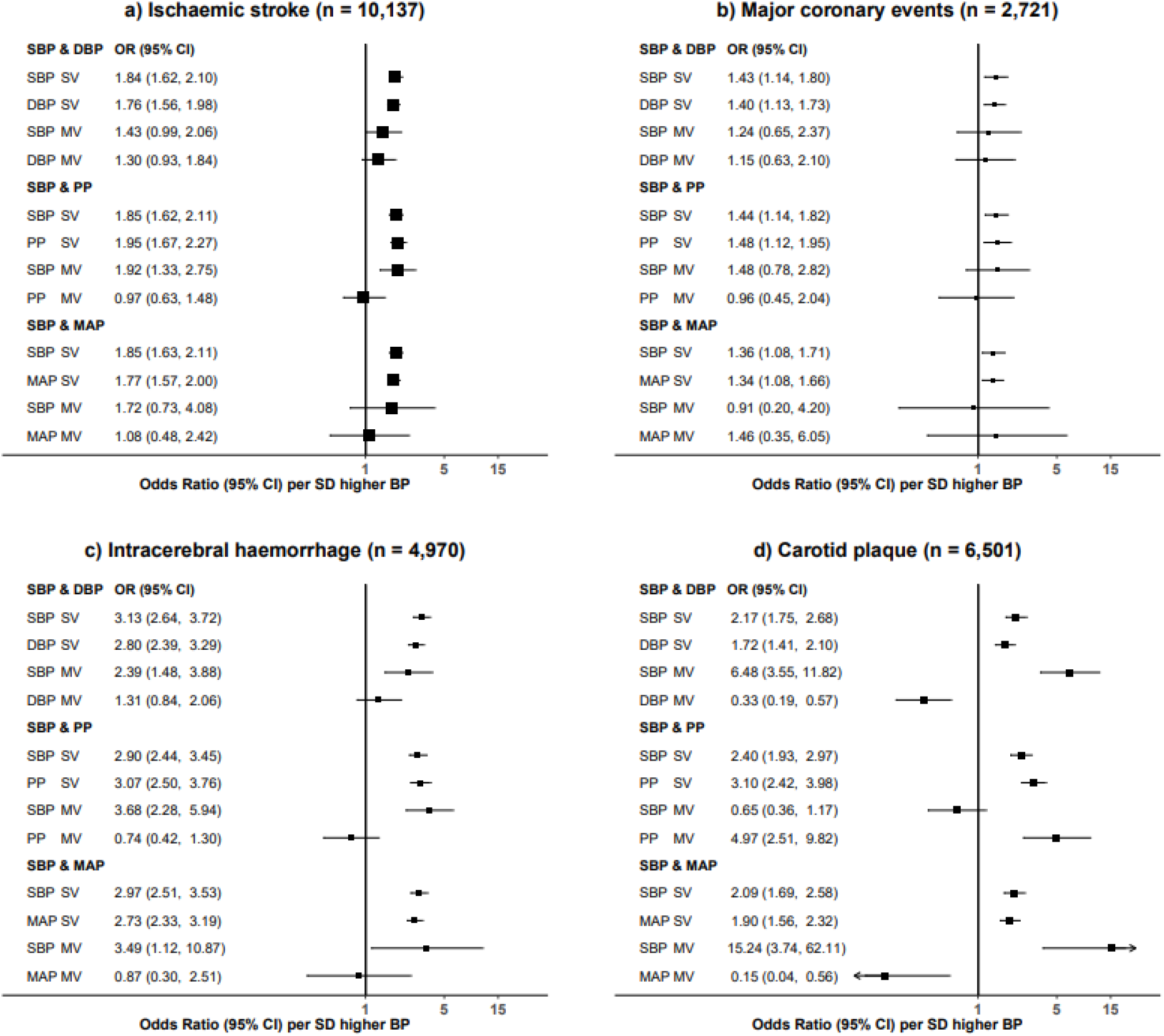
Multivariable Mendelian randomisation of blood pressure traits with risk of major cardiovascular diseases and subclinical atherosclerosis. Effects are shown as odds ratios (95% CI) of disease risk per 1-SD higher blood pressure. Loci from two traits were merged and in cases where more than one variant was available in the locus, the variant with the lowest p-value was included as the instrument when constructing the polygenic score. SBP indicates systolic blood pressure; DBP, diastolic blood pressure; PP, pulse pressure; MAP, mean arterial pressure; SV, single variable MR; MV, multi-variable MR.

For IS, mutual adjustment of SBP and DBP resulted in partial attenuation of their effects observed in univariate MR, giving ORs of 1.38 [0.96-1.99] and 1.33 [0.95-1.87] per 1-SD higher BP, respectively, while neither PP nor MAP contributed independently to IS risk after adjusting for SBP (**Figure 4a**). A similar overall pattern was observed for MCE, although there was an insufficient number of cases to reliably distinguish which BP traits were independent risk factors (**Figure 4b**). However, the pattern of causal relationships was different for ICH – only SBP was independently associated with increased risk of ICH, with each of DBP, PP, and MAP being attenuated to the null after adjustment for SBP (**Figure 4c**); in these models, adjusted ORs for SBP were between 2.30 [1.42-3.72] and 3.69 [2.31-5.94], consistent with the estimate from univariate MR. MVMR results for CP were clearly different from those for the other disease outcomes. Mutual adjustment of PP and SBP attenuated the association with SBP to the null, while that with PP was not appreciably unaffected (**Figure 4d**). Analyses of SBP with each of DBP or MAP gave unstable models in which DBP and MAP appeared to have protective effects, but further models in which PP was mutually adjusted for DBP or for MAP yielded consistent results in which effects of DBP and MAP were attenuated to the null, while PP retained strong association with risk of CP with ORs similar to those from univariate MR (**Supplementary Figures 13**).

To ensure MVMR results were not biased by selection of individual variants more strongly associated with one of a pair of traits, we performed additional MVMR analyses using two further sets of GSs that selected variants from a locus according to the strength of their association with one or other of the traits being analysed; these GSs yielded MVMR results very similar to those from the main analyses (**Supplementary Figures 14-15**). We also attempted MVMR including all 4 traits in a single model, but these analyses invariably yielded unstable estimates.

## Discussion

In this large study of the genomics of four BP measures in Chinese adults, we identified a total of 128 genetic loci significantly associated with BP, of which 36 had not previously been identified as associated with BP in East Asians, 13 being newly associated with BP in any ancestry. We identified 81 new trait-locus associations, of which a large proportion were for MAP, likely reflecting that MAP has been less widely investigated in previous studies. About half of the novel associations identified in Chinese were replicated in analyses of Japanese and/or European adults. For all BP traits, narrow sense heritability and effect sizes of relevant SNPs were greater in Chinese compared with other populations, especially Japanese. GSs derived from these loci explained 2.6% to 3.3% variance of different BP traits. In MR analyses, all four BP traits showed highly significant associations with risks of different CVD types, with comparable per 1-SD risk estimates which were greater for ICH than for IS and MCE. For carotid plaque, however, PP was more strongly associated than other BP traits.

BMI is a strong risk factor for BP in both observational and MR studies ^33-35^. Previous GWAS of BP have variously included or omitted adjustment for BMI, so to improve comparability we performed GWAS both with and without such adjustment. We found that adjustment for BMI had little impact on SNP effect sizes (with the exception of the *FTO* locus), and did not affect the significance of association for the majority of BP loci. Indeed, adjustment for BMI increased the number of genomic regions associated with different BP phenotypes. We infer that, although BMI overall impacts on BP, the principal biological pathways associated with adiposity do not directly regulate or impact on BP.

More than 2,000 associations with various BP phenotypes have previously been identified.^21-23,25-29,36^ The loci corresponding to these associations overlap with signals for many other traits (e.g. obesity, lipids) and cardiovascular disease outcomes.^22^, potentially informing development of drugs for treatment of hypertension. Pathway analyses have identified enrichment of BP signals at loci with roles in the structure of arterial walls, and have indicated that TGF-β and Notch signalling pathways play an important role in BP regulation^22,26,36^. Previous studies have been primarily conducted in individuals of European ancestry, raising questions about the relevance of these findings to other ancestry populations, but more recent analyses have improved our understanding of the genetic architecture of BP in other ancestries, and have established that many BP associations are replicated in diverse populations^21,23,28,29^. Nevertheless, we have demonstrated that large studies in non-European populations provide continuing opportunities for discovery of novel genetic associations with BP. In the present study, we identified 13 novel BP loci, despite having a sample size that was approximately one-tenth that of previous studies of European individuals ^22,26,36^ and similar to those of previous studies of East-Asian populations.^21,23,28,29^

SBP and DBP are the most widely used measures of blood pressure in clinical practice. Although both PP and MAP are directly derived from SBP and DBP, they reflect discrete aspects of biology. PP (calculated as the difference between SBP and DBP) potentially reflects differences in cardiac structure or wall thickness caused by sustained hypertension, valvular regurgitation, age-related aortic stiffness, severe iron deficiency anaemia, or hyperthyroidism.^37,38^ MAP (less often included in genetic analyses) represents the mean pressure during a cardiac cycle and is influenced by cardiac output, systemic vascular resistance, volume and viscosity of circulating blood, and the elasticity of vessel walls.^39^ Consistent with previous reports in different ancestries^40,41^, we found strong genetic correlation between these different BP traits. Accordingly, the majority (∼60%) of BP loci in the present analyses were associated with multiple BP phenotypes. Nevertheless, 24 loci would not have been identified by considering only SBP and DBP, highlighting the value of conducting analyses of multiple related traits.

Previous GWAS of BP have highlighted the highly heritable nature of BP, with estimates of “narrow sense” heritability (due to additive genetic effects) for SBP and DBP in the range of 17-52%.^42^ We observed systematically lower heritability for all traits in European and Japanese populations compared to Chinese. In particular, heritabilities in BBJ were less than half those in CKB. By contrast, LD score estimates of *h*^2^ for SBP and DBP reported for the Taiwan Biobank (TWB)^29^ were similar to those for CKB. Although our estimates of heritability based on CKB, BBJ, and ICBP summary statistics using LD score regression were lower (**Table 1**), this likely reflects that these estimates reflect common variation only; substantially higher *h*^2^ estimates (19-21%) were reported by ICBP for UK Biobank using REML as implemented in BOLT-LMM^22^, and we found comparable increases in heritability estimates in CKB (20-27%) when using REML (**Supplementary Table 20**).

In addition to lower heritabilities compared to CKB, variant effect size estimates were also substantially lower in both ICBP and BBJ (**Figure 2, Supplementary Figure 11**). Although several previous studies have also examined differences in heritability across populations,^21,26^ we are unaware of studies that have reported systematic differences in variant effect sizes on the scale observed. Nevertheless, despite these large differences in estimated heritability and effect sizes, we found strong genetic overlap of BP traits across ancestries and between East Asian populations, consistent with previous findings for several other diseases and complex traits^32,43,44^. This suggests that the observed differences in heritability do not reflect major differences in the genetic architecture of BP traits. Rather, we suggest that other external factors may modify the extent to which genetic variation influences BP, leading to parallel changes in variant effect and, consequently, heritability.

There are multiple differences between the CKB, BBJ and ICBP populations that might impact on effect size and heritability. In particular, CKB is a population-based cohort (as is TWB), while BBJ is based on a combination of different hospital-based disease cohorts, who may have different BP characteristics from healthy adults. Furthermore, by contrast with Japanese and European-ancestry individuals, hypertension is still poorly detected, treated and controlled in China^5^. A nationally representative sample of ∼450K individuals in China reported that around half of hypertensive patients did not take prescribed BP-lowering medications, and BP was controlled for only one in six of those treated^6^; similarly, in CKB BP was successfully controlled only in approximately 5% of participants with hypertension.^2^ Thus, although all analyses included adjustment for use of blood pressure-lowering medication, the findings of the present study may reflect the impact of performing analyses in largely untreated populations. Another possibility is that differences in lifestyle, including dietary patterns, may contribute to differences in measured heritability and variant effect sizes. Sodium intake is associated with hypertension^16,45^ in addition to higher risks of chronic diseases^17-19,46^ and salt intake in China has been estimated to be among the highest in the world.^46^

Although observational associations of higher levels of BP with increased CVD risk are well established, the strong correlation between individual BP traits makes it very difficult to establish the independent causal relevance of individual BP traits. Similarly, previous MR studies have demonstrated apparent causal effects of multiple BP traits on different CVD types^47-53^. Consistent with these, using strong genetic instruments derived from genome-wide significant associations from our GWAS, all four BP traits showed similar per-SD associations with risk of different CVD types in Chinese, with SBP being associated with stronger risk than DBP. Notably, however, when expressed per unit change in SBP (i.e. per 5 mmHg), SBP showed the weakest associations with CVD outcomes, while DBP had the strongest association. The odds ratios from these analyses were consistent with those from observational analyses in CKB.^2^ By contrast, for CP the strongest association was consistently observed for PP, suggesting a possibly different causal relevance for CP than for CVD outcomes.

Although suggestive, such MR analyses do not provide conclusive evidence about which traits have a true causal relationship with different CVD-related outcomes. The strong correlations between variant effect sizes for different BP measures (**Supplementary Figure 9**) suggest that a GS for one trait is also an effective instrument for another and cannot completely discriminate between them. For this reason we employed MVMR, in which the association with one instrument is estimated conditional on a second instrument for a different trait, but with both instruments using identical genetic variants. We observed mutual partial attenuation by SBP and DBP for risk of IS (and also for MCE, although this analysis was less well-powered), suggesting that SBP and DBP each independently contributed to disease risk Very few previous studies have assessed the independent effects of different BP traits, and none have had sufficient power to investigate the independent effects of BP traits on ICH risk. The associations of DBP, MAP, and PP instruments with ICH were completely attenuated to the null by inclusion of an SBP instrument, whose association was largely unaffected in the joint analysis. Thus, we conclude that BP association with ICH risk is almost entirely explained by associations with SBP. Similarly, we have identified a specific causal association of PP with CP, which was unaffected by inclusion of instruments for other BP traits whose associations were attenuated to the null by inclusion of a PP instrument. This is consistent with previous observational studies which reported strong associations of PP with CP, believed to relate to arterial remodelling in response to turbulence at the bifurcation of the common carotid arteries, resulting in development of carotid plaques^54,55^.

Previous MVMR of BP traits have been limited to 2-sample studies of SBP and DBP, based on summary statistics for ischaemic CVD in populations of European ancestry,^47,56^ in whom the proportions of different subtypes of CVD differ substantially from those in CKB (e.g. much larger contributions of cardioembolic stroke).^57^ Both found that affects due to DBP were attenuated more strongly than those from SBP, but both studies had wide confidence intervals and are consistent with our findings that both SBP and DBP contribute to IS risk. Importantly, the ICH and CP analyses demonstrate that we were able to reliably distinguish specific independent effects of individual BP traits on disease risk. The absence of such a finding in the better-powered IS analysis (with approximately twice as many cases as ICH or CP), strongly suggests that the mutual attenuation by SBP and DBP instruments for IS reflects a genuine causal contribution by both traits.

The present study has several strengths, including prospective study design, standardised data collection across the entire study population, comprehensive capture of disease outcomes with validation of diagnoses through retrieval of medical notes including details of imaging and other investigations, large sample size in an understudied population with different prevalences of disease subtypes, and availability of individual participant data. We were able to perform well-powered GWAS with replication of novel variants in independent populations and, in addition to comparisons of GWAS of multiple BP traits, we performed systematic assessment of variant effect sizes and heritability across different ancestries. This enabled us to construct strong genetic instruments that could be used for MR and MVMR analyses of different BP traits with several CVD outcomes. Nevertheless, the study had several limitations: GWAS discovery was conducted within study population used for MR, but restricting instruments to genome-wide significant loci limited the potential for weak instrument bias; there remained a degree of collinearity between genetic instruments, in some analyses leading to unstable MVMR estimates, but the use of 4 different traits meant that these could be identified as occurring when no causal trait was included; and there remained the possibility of bias in MR analyses due to horizontal pleiotropy, but consistent results across three different methods used to select instruments for MVMR strongly suggests this was not a major factor.

Overall, this large genetic study of Chinese adults identified a total of 128 genetic loci associated with BP traits, of which 13 had not been previously reported. Across three populations, there were substantial differences in heritability and variant effects for all BP traits. Although all four BP traits showed highly significant associations with risks of different CVD types in MR analyses, their causal relevance differed, with SBP causal for ICH, PP independently associated with CP, and both SBP and DBP making independent contributions to IS risk. The findings of this study should enhance our understanding of the genetic architecture of BP traits in different ancestries, and of the biological processes responsible for BP-associated increases in CVD risk.

## Online methods

### Study population

The China Kadoorie Biobank (CKB) is a prospective study of >513,000 Chinese individuals aged 30-79 years recruited from 10 geographically defined regions during 2004-08. CKB recruitment and study design has been described in detail elsewhere^58-60^. In brief, the baseline survey collected extensive information about participants’ socio-demographic status, lifestyle factors, environmental exposures, medical history (including medication use) and physical characteristics, through an interviewer-administered questionnaire and physical measurements (**Supplementary Table 1**). Data on disease outcomes were identified from claims to the national health insurance system and from local chronic disease registries and disease surveillance point system death registries. All events were ICD-10 coded by trained staff blinded to baseline information. Active annual follow-up through local residential and administrative records confirmed the participant’s vital status. Each participant also provided a blood sample for long-term storage. After the baseline survey, 5% of randomly selected surviving participants were re-surveyed periodically. In the 2013-14 resurvey, as well as repeating measurements made at baseline, carotid ultrasound examination was performed to assess cIMT and carotid plaque. The presence of carotid artery plaque was defined as focal thickening or protrusion from the artery wall into the lumen with carotid intima-media thickness >1.5 mm^61^. All participants provided written informed consent at each survey visit, allowing access to their medical records and long-term storage of biosamples for future unspecified medical research purposes, without any feedback of results to the individuals concerned. Ethical approval was obtained from the Oxford Tropical Research Ethics Committee, the Ethical Review Committees of the Chinese Centre for Disease Control and Prevention, Chinese Academy of Medical Sciences, and the Institutional Review Board (IRB) at Peking University.

### Blood pressure measurement

At baseline, BP was measured twice with a 5-min interval between measurements using Omron UA-779 digital sphygmomanometers (A&D Instruments; Abingdon, UK) that were regularly maintenanced and calibrated. A third BP measurement was recorded only if the difference between the previous two SBP measurements was greater than 10 mmHg. The mean of the last two readings was recorded and used for analyses.^2^ Participants who reported receiving antihypertensive medication at baseline had their recorded BP adjusted by adding a constant value for SBP (15mmHg) and DBP (10mmHg). Using these adjusted values, PP was calculated as SBP - DBP and MAP was calculated as (2 x DBP + SBP)/3.

BP phenotypes for GWAS were derived from the whole CKB cohort of 513,214 individuals, before exclusions (**Supplementary Figure 1**). Two individuals with missing BMI measurements and 371 individuals with BP measurements greater than 5 SDs from the population mean were excluded. After linear regression of each BP trait on age, age^2^, sex, CKB study region, mean monthly outdoor temperature (temperatures below 5C were set to 5C),^62^ and BMI, an additional 321 individuals were excluded with residuals greater than 5 standard deviations from the mean for one or more measures of BP. The regressions were repeated for the remaining 100,453 participants, with/without adjustment for BMI, and residuals from the regression for these participants were used in association analyses.

### Genotyping and SNP quality control

Genotyping and quality control has been described in detail elsewhere^63^. After exclusions of samples with low call rate (< 95%), high heterozygosity (3SD above the mean), sex mismatch, ancestry outliers, XY aneuploidy, bad linkage, and missing/withdrawn consent, genotyping data were available for 100,706 participants. After imputation into the 1000 Genomes Phase 3 reference, variants with MAF < 0.005 and imputation accuracy (*r*^2^_INFO_) < 0.3 were excluded. For analyses of each CKB geographic region separately, an additional 6,096 individuals with non-local ancestry were excluded^63^. All genomic locations are specified using the GRCh37 build.

### Vascular outcomes

Causal relationships were assessed between BP phenotypes and incident vascular events, in which cases were defined according to the first cardiovascular event (multiple events on the same day were excluded): 10,137 ischaemic stroke cases (IS, ICD-10: I63); 4,970 intracerebral haemorrhage cases (ICH, ICD-10: I61); and 2,721 major coronary events (MCE) comprising myocardial infarction (MI, ICD-10: I21-I23) or fatal ischaemic heart disease (IHD, ICD-10: I20-I25). 72,587 common controls with no cardiovascular events were drawn from a population representative subset of genotyped participants.^63^ For analyses of carotid plaque (CP, 6,501 cases), controls were all other genotyped second resurvey participants with available carotid ultrasound data.

### GWAS

Genome-wide association analyses using residualised BP phenotypes for 100,453 genotyped participants were conducted using *BOLT-LMM* version 2.3.2^64^, with array version as covariate. As a sensitivity analysis, GWAS were performed in each CKB region separately and summary statistics were meta-analysed using an inverse-variance-weighted fixed-effect model in *METAL*^65^.

We estimated the genomic inflation (*λ*_GC_) of CKB BP GWAS to range from 1.20 to 1.31 (mean *λ*_GC_ = 1.261). Using pre-computed LD scores for East Asian and European ancestries (https://data.broadinstitute.org/alkesgroup/LDSCORE/), LD score regression intercepts (standard errors) for BMI-adjusted models were 1.056 (0.010), 1.054 (0.011), 1.039 (0.009), and 1.060 (0.011) for SBP, DBP, PP, and MAP, respectively. These were substantially smaller than the mean χ^2^ and were comparable with those for other studies (**Supplementary Table 21**), indicating no substantial inflation except for that due to the polygenic nature of BP traits. Assessment of genomic inflation for BMI-unadjusted analyses was similar.

### Heritability and genetic correlation

Estimates of narrow-sense heritability and between-trait genetic correlation used LD score regression applied to summary statistics from this study, BBJ^21^, and ICBP^22^ for ∼1M HapMap3 SNPs with MAF>0.05, INFO>30 and ꭓ^2^<30 and excluding variants in the HLA region (chr6:21Mb-41Mb)^66^. LD score regression was also used for additional comparisons between CKB and BBJ. Cross-ancestry/cross-population genetic correlations for each trait between studies were estimated using *Popcorn*^32^ and cross-population LD scores for variants with MAF > 0.01 in the East Asian populations of the 1000 Genomes Project Phase 3^67^.

### Locus identification

Genomic regions defined by genome-wide significant variants (P<5×10^−8^) were defined by LD-based clumping in *PLINK*^68^ (initial window +/-10Mbp, P<0.05, LD r^2^>0.05) using an internal LD reference of 10,000 unrelated CKB participants with imputed genotype probabilities converted to “best-guess” genotypes. We observed that, for three loci, the lead variant lay out outside the reported clumped range (all variants in LD lay either proximal or distal to the lead variant); in this instances we extended the clumped region to include the lead SNP. All locus boundaries were then extended by an additional 1Kbp in each direction. Overlapping loci were merged and the variant with the lowest *P*-value was identified as the sentinel. Conditional analyses to identify independent associations within a locus were performed using the stepwise model selection procedure in *GCTA*^69^. For comparisons of loci across traits, all overlapping loci identified for any trait were merged into single larger genomic regions which were then treated as single loci.

Locus novelty was assessed by testing whether any previous lead variant from previous studies of BP^21-23,25,28,29,36^, or reported in the HGRI-EBI GWAS catalog^30^ (accessed on 14/06/2022), lay within the locus. Assessment of novelty in East Asians used only lead variants from the relevant studies^21,23,28,29^. Replication of 81 novel associations was conducted using an independent sample from BioBank Japan (BBJ, n_max_=133,567) or *in silico* using existing publically available ICBP summary statistics^22^. In the Japanese sample, phenotype residuals were obtained after adjusting for sex, age, age^2^, first 10 principal components, disease status for the 47 target diseases in the BBJ, and with or without adjustment for BMI (**Supplementary Figure 16**). Replication was assessed at 5% false discovery rate.

### Comparison of SNP effects across BP traits and ancestries

Comparisons of variant effects, between BMI-adjusted and BMI-unadjusted models, between traits, and between populations, were each assessed used Deming regression^70^ as implemented in R (https://cran.r-project.org/package=deming). For comparisons between pairs of populations with the third excluded, the variants compared were lead variants as reported by the excluded study. BBJ GWAS summary statistics^23^ for all phenotypes were downloaded from http://jenger.riken.jp/en/result. ICBP GWAS summary statistics were downloaded from http://biota.osc.ox.ac.uk) and from https://ftp.ncbi.nlm.nih.gov/dbgap/studies/phs000585/analyses. BBJ summary statistics (expressed in SD units) were converted to mmHg using the phenotype-specific SD (**Supplementary Table 2**).

### Mendelian randomisation

MR was conducted using genetic scores constructed from the sentinel variants at each associated locus in the BMI-adjusted analyses. For univariate MR, the score for each trait was the sum of the dosages of risk alleles for that trait, weighted by the effect size from the corresponding GWAS. For MVMR with a pair of traits, a single sentinel variant was selected for each locus associated with either of the traits (overlapping loci were merged and treated as a single locus); where there was a choice of two sentinel variants (one for each trait), the variant with the lowest *P*-value was selected. The score for each trait was the sum of the dosage of risk alleles for the selected variants, weighted by the effect sizes for those variants from the corresponding GWAS. To assess potential bias due to greater power in the GWAS for one of the pair of traits, two alternative scores were derived in which selected variants were always those most strongly associated with one of the traits being analysed.

Linear regression of each BP trait on its GS, with BMI, sex, age, age^2^, and regional principal components as covariates, was performed to derive the coefficient for the GS; this was used to scale individual GS values to derive genetically-predicted BP traits. Associations of these genetically-predicted exposures with binary vascular events were then assessed using logistic regression, with adjustment for sex, age, and age^2^. For MVMR, pairs of genetically-predicted BP traits were mutually adjusted to give causal estimates for each trait independent of the other. Both MR and MVMR were performed separately in each CKB recruitment region, and the estimates from each region were combined using inverse-variance weighted fixed effect meta-analysis.

For each GS, F-statistic and variance explained (estimated as the partial *r*^2^ as the proportional reduction of the error sum of squares (SSE) of a full regression model including the GS compared to a reduced model without the GS) were as calculated by the R function anova(). For calculation of conditional F-statistic and r^2^ in MVMR, the models included the GS for the second trait in the analysis.

## Data Availability

The China Kadoorie Biobank (CKB) is a global resource for the investigation of lifestyle, environmental, blood biochemical and genetic factors as determinants of common diseases. The CKB study group is committed to making the cohort data available to the s cientific community in China, the UK and worldwide to advance knowledge about the causes, prevention and treatment of disease. Full details of what data is currently available to open access users and how to apply for it, visit: http://www.ckbiobank.org/site/Data+Access. Researchers who are interested in obtaining the raw data from the China Kadoorie Biobank study that underlines this paper should contact. A research proposal will be requested to ensure that any analysis is performed by bona fide researchers and - where data is not currently available to open access researchers - is restricted to the topic covered in this paper.

## Acknowledgements

The chief acknowledgment is to the participants, the project staff, and the China National Centre for Disease Control and Prevention (CDC) and its regional offices for assisting with the fieldwork. We thank Judith Mackay in Hong Kong; Yu Wang, Gonghuan Yang, Zhengfu Qiang, Lin Feng, Maigeng Zhou, Wenhua Zhao, Yan Zhang and Zheng Bian in China CDC; Lingzhi Kong, Xiucheng Yu, and Kun Li in the Chinese Ministry of Health; and Garry Lancaster, Sarah Clark, Martin Radley, Mike Hill, Hongchao Pan, and Jill Boreham in the CTSU, Oxford, for assisting with the design, planning, organization, and conduct of the study. Members of the China Kadoorie Collaborative Group are listed in the supplementary material.

## Funding

The CKB baseline survey and the first re-survey were supported by the Kadoorie Charitable Foundation in Hong Kong. The long-term follow-up has been supported by Wellcome grants to Oxford University (212946/Z/18/Z, 202922/Z/16/Z, 104085/Z/14/Z, 088158/Z/09/Z) and grants from the National Key Research and Development Program of China (2016YFC0900500, 2016YFC0900501, 2016YFC0900504, 2016YFC1303904) and from the National Natural Science Foundation of China (91843302). DNA extraction and genotyping was supported by grants from GlaxoSmithKline and the UK Medical Research Council (MC-PC-13049, MC-PC-14135). The UK Medical Research Council (MC_UU_00017/1,MC_UU_12026/2 MC_U137686851), Cancer Research UK (C16077/A29186; C500/A16896) and the British Heart Foundation (CH/1996001/9454) provide core funding to the Clinical Trial Service Unit and Epidemiological Studies Unit at Oxford University for the project. PKI is supported by an Early Career Research Fellowship from the Nuffield Department of Population Health, University of Oxford.

Computation used the Oxford Biomedical Research Computing (BMRC) facility, a joint development between the Wellcome Centre for Human Genetics and the Big Data Institute supported by Health Data Research UK and the NIHR Oxford Biomedical Research Centre. The views expressed are those of the author(s) and not necessarily those of the NHS, the NIHR or the Department of Health.

## Authors’ contributions

AP, WG, and KL analysed the data. AP drafted the manuscript. AP, WG, KL, IYM, LL, ZC and RGW contributed to the conception of this paper, interpretation of the results and the revision of manuscript. RGW, LL and ZC designed the study. KL, MY, YG, CM, JL, CY, DA, DSV, LL, ZC, IYM, and RGW, contributed to data acquisition. All authors critically reviewed the manuscript and approved the final submission.

## Conflict of interest

The authors declare that they have no competing interests.

## Data availability statement

The China Kadoorie Biobank (CKB) is a global resource for the investigation of lifestyle, environmental, blood biochemical and genetic factors as determinants of common diseases. The CKB study group is committed to making the cohort data available to the s cientific community in China, the UK and worldwide to advance knowledge about the causes, prevention and treatment of disease. Full details of what data is currently available to open access users and how to apply for it, visit: http://www.ckbiobank.org/site/Data+Access.

Researchers who are interested in obtaining the raw data from the China Kadoorie Biobank study that underlines this paper should contact. A research proposal will be requested to ensure that any analysis is performed by bona fide researchers and - where data is not currently available to open access researchers - is restricted to the topic covered in this paper.

## Ethics statement

Ethical approval was obtained from the Ethical Review Committee of the Chinese Centre for Disease Control and Prevention (Beijing, China, 005/2004) and the Oxford Tropical Research Ethics Committee, University of Oxford (UK, 025-04), and all participants provided written informed consent.

## Open access statement

This research was funded in whole, or in part, by the Wellcome Trust [212946/Z/18/Z, 202922/Z/16/Z, 104085/Z/14/Z, 088158/Z/09/Z]. For the purpose of Open Access, the author has applied a CC-BY public copyright licence to any Author Accepted Manuscript version arising from this submission.

## Notes

### Competing Interest Statement

Wei Gan is now an employee of Novo Nordisk Research Centre Oxford Ltd

