## Supplementary figures 1-16 for "Causal relevance of different blood pressure traits on risk of cardiovascular diseases: GWAS and Mendelian randomisation in 100,000 Chinese adults": Supplementary_Figure5.docx

**Supplementary Figure 5. Manhattan plots and MAF-stratified quantile-quantile (Q-Q) plots for association with mean arterial pressure**. Genome-wide significant trait-specific loci are highlighted in blue. Novel loci are highlighted in red.

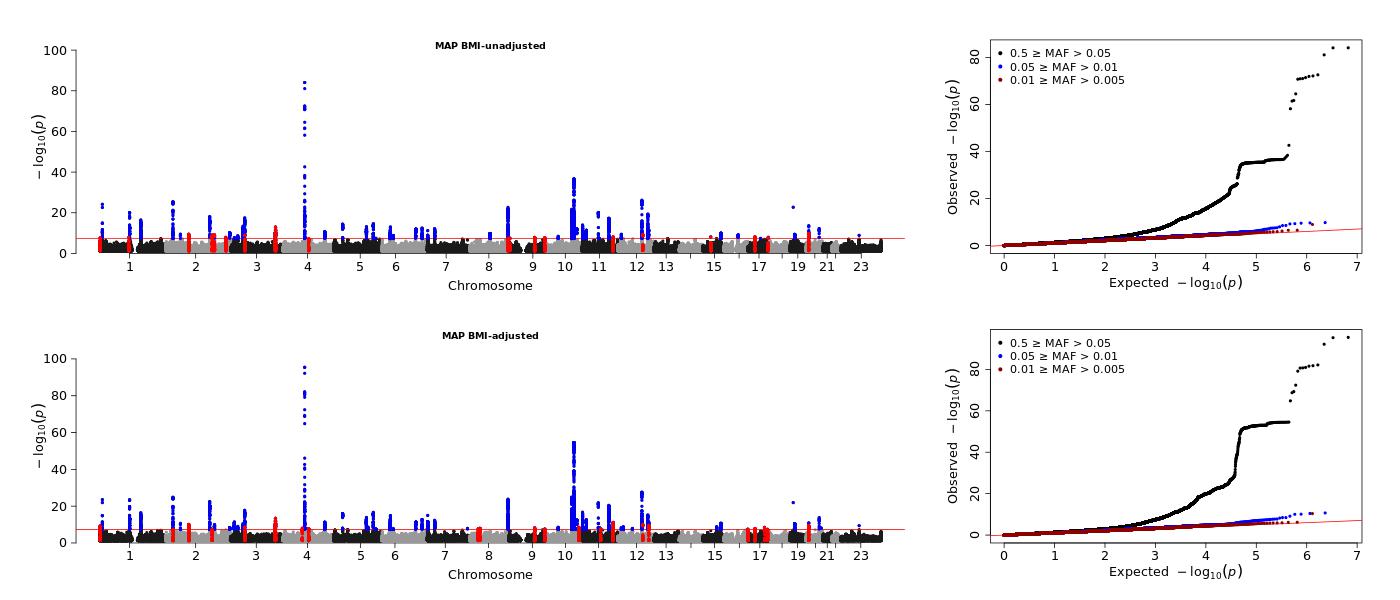
