## Supplementary figures 1-16 for "Causal relevance of different blood pressure traits on risk of cardiovascular diseases: GWAS and Mendelian randomisation in 100,000 Chinese adults": Supplementary_Figure7.docx

**Supplementary Figure 7. Overlap of associations across BMI-unadjusted blood pressure traits.** (a) Venn diagram of associations across BMI-adjusted blood pressure traits. The numbers in brackets indicate the total number of loci associated with that BP phenotype. (b) Genetic correlation between 4 traits as assessed by LD score regression.

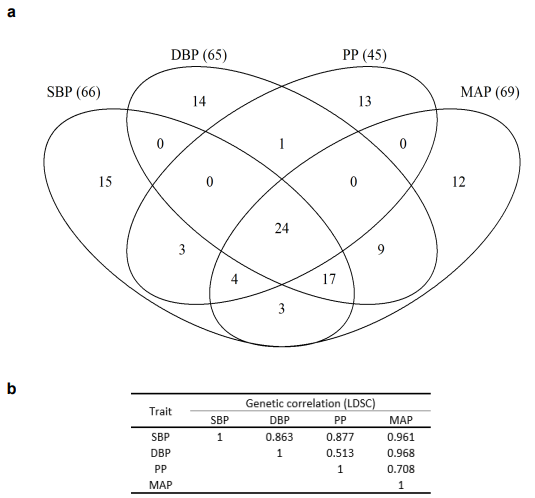
