## Supplementary figures 1-16 for "Causal relevance of different blood pressure traits on risk of cardiovascular diseases: GWAS and Mendelian randomisation in 100,000 Chinese adults": Supplementary_Figure8.docx

**Supplementary Figure 8. Overlap of novel associations across BMI-adjusted blood pressure traits.** The numbers in brackets indicate the number of novel loci associated with that BP phenotype. SBP indicates systolic blood pressure; DBP, diastolic blood pressure; PP, pulse pressure; MAP, mean arterial pressure.

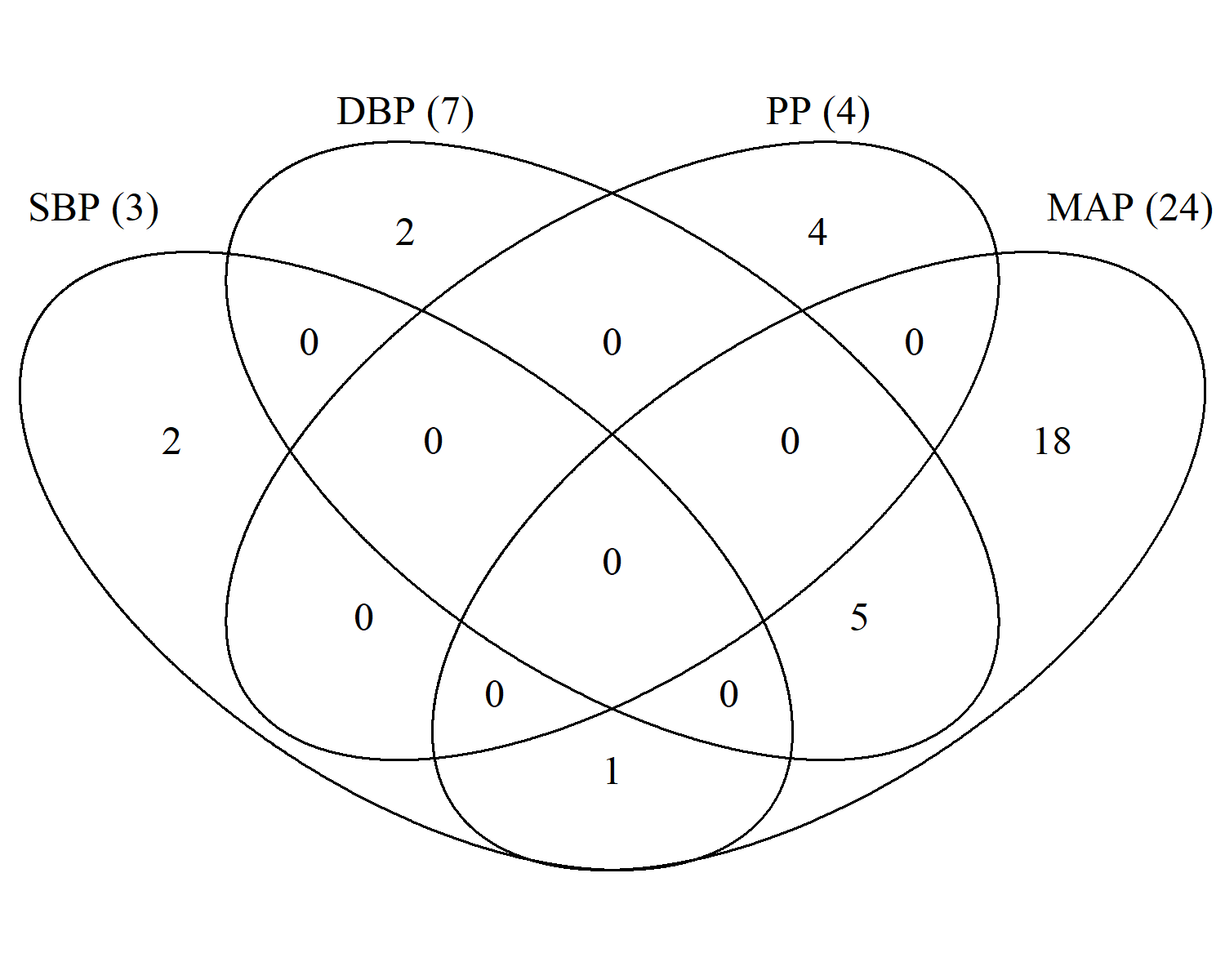
