## Supplementary figures 1-16 for "Causal relevance of different blood pressure traits on risk of cardiovascular diseases: GWAS and Mendelian randomisation in 100,000 Chinese adults": Supplementary_Figure9.docx

**Supplementary Figure 9. Comparison of CKB trait effect sizes for SNPs associated with BMI-adjusted blood pressure traits.** Effect sizes for SNPs associated with each BP phenotype (in columns) were compared with their effect sizes for the remaining BP phenotypes (in rows). Dashed diagonal lines represent identity (y = x). Solid lines are derived from Deming regression, forced through the origin. SBP, systolic blood pressure; DBP, diastolic blood pressure; PP, pulse pressure; MAP, mean arterial pressure

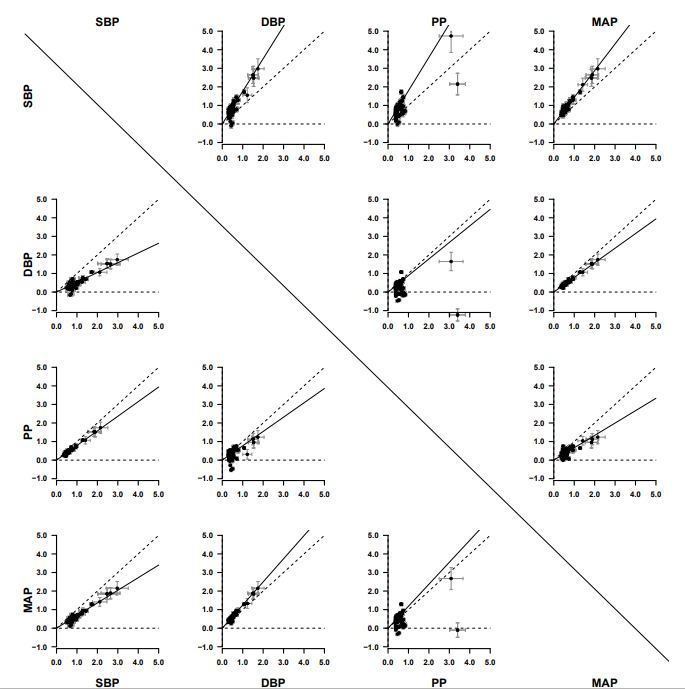
