## Supplementary figures 1-16 for "Causal relevance of different blood pressure traits on risk of cardiovascular diseases: GWAS and Mendelian randomisation in 100,000 Chinese adults": Supplementary_Figure10.docx

**Supplementary Figure 10. Comparison of effect sizes for novel blood pressure associations in CKB and BBJ.** SNP effects are shown using the mmHg scale. The dashed diagonal line is the identity line (y = x). The solid line is the Deming regression line forced through the origin.

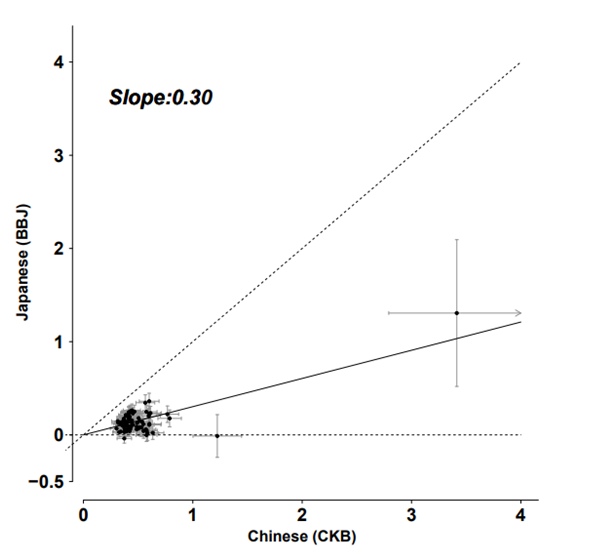
