## Supplementary figures 1-16 for "Causal relevance of different blood pressure traits on risk of cardiovascular diseases: GWAS and Mendelian randomisation in 100,000 Chinese adults": Supplementary_Figure11.docx

**Supplementary Figure 11. Comparisons of variant effect sizes of blood pressure traits between BBJ (BMI-unadjusted), ICBP (BMI-adjusted), and CKB (BMI-unadjusted and BMI-adjusted) cohorts.** Comparisons of urban (a) and rural (b) regions of CKB with ICBP and BBJ used variants identified in BBJ and ICBP, respectively. Variant per-allele effects are shown on the mmHg scale. The dashed diagonal line is the identity line (y = x). The solid line is the Deming regression line forced through the origin.

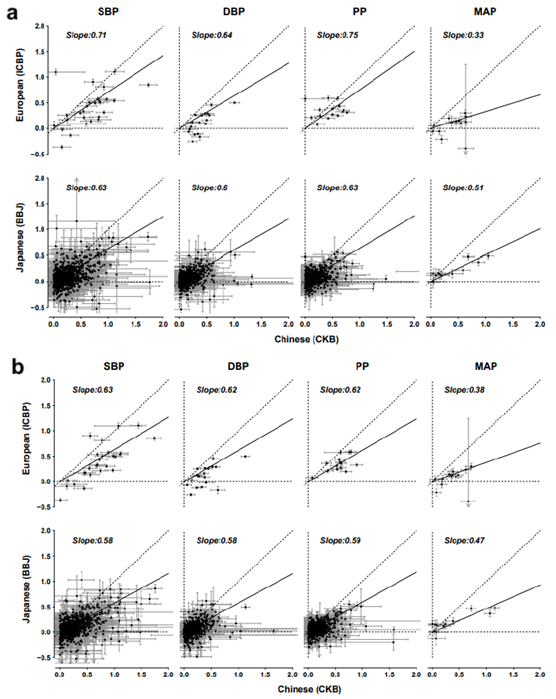
