## Supplementary figures 1-16 for "Causal relevance of different blood pressure traits on risk of cardiovascular diseases: GWAS and Mendelian randomisation in 100,000 Chinese adults": Supplementary_Figure12.docx

**Supplementary Figure 12. Region-specific heritability in CKB.** Heritabilities were estimated using *BOLT-*REML**.** SBP indicates systolic blood pressure; DBP, diastolic blood pressure; PP, pulse pressure; MAP, mean arterial pressure; BMI, body mass index.

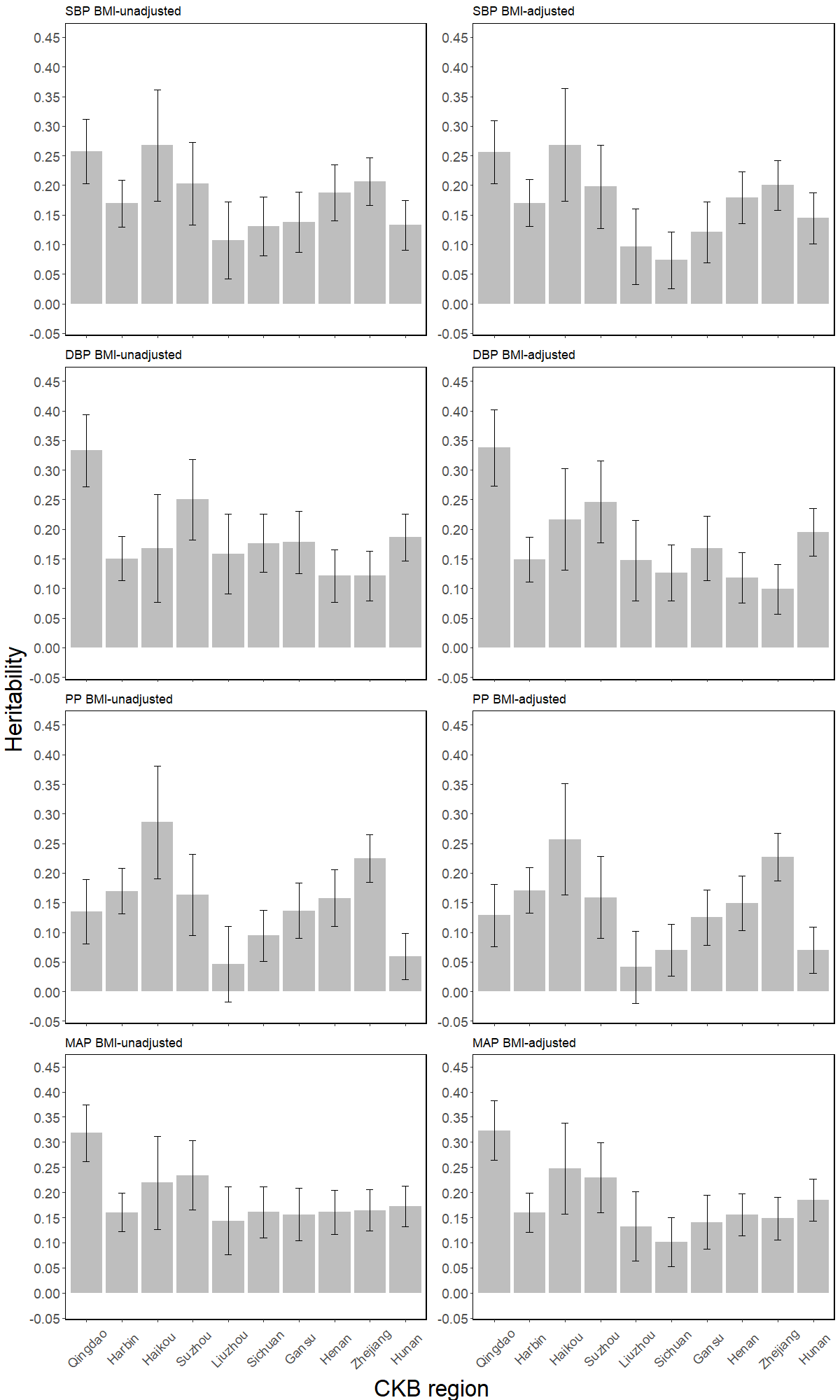
