## Supplementary figures 1-16 for "Causal relevance of different blood pressure traits on risk of cardiovascular diseases: GWAS and Mendelian randomisation in 100,000 Chinese adults": Supplementary_Figure13.docx

**Supplementary Figure 13. Multivariable Mendelian randomisation of blood pressure with major cardiovascular diseases and subclinical atherosclerosis**. Effects are shown as odds ratios (95% CI) of disease risk per 1 SD higher blood. Loci from two traits were merged and in cases where more than one variant was available in the locus, the variant with the lowest p-value was included as the instrument when constructing the polygenic score. SBP indicates systolic blood pressure; DBP, diastolic blood pressure; PP, pulse pressure; MAP, mean arterial pressure; SV, single variable MR; MV, multi-variable MR.

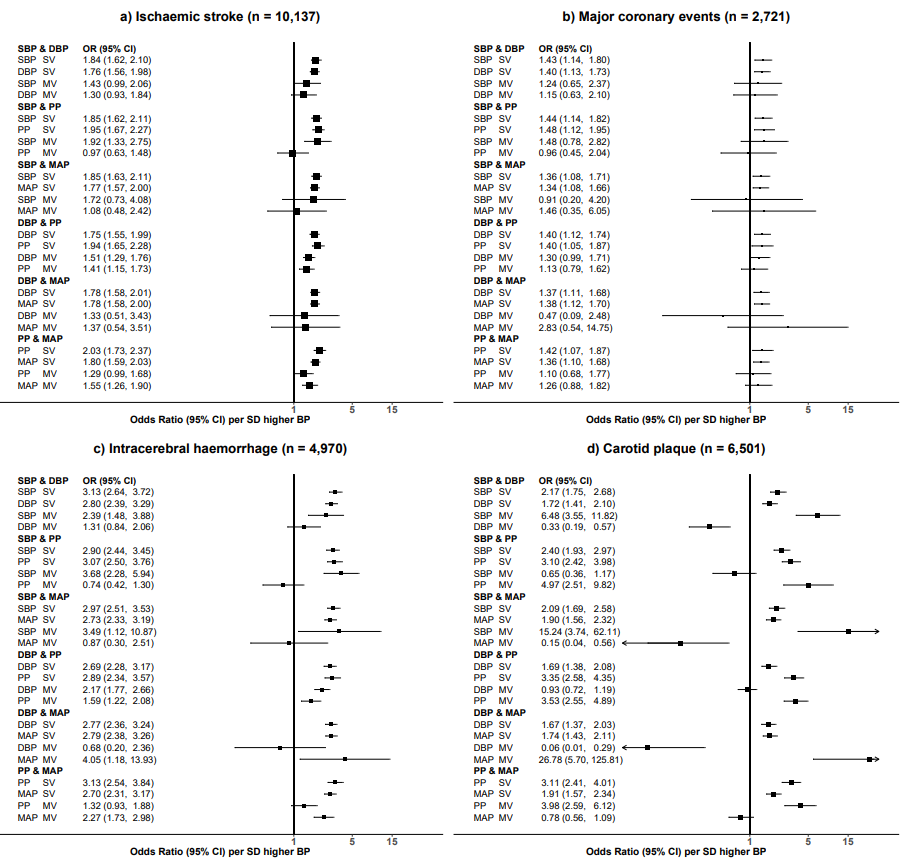
