## Supplementary figures 1-16 for "Causal relevance of different blood pressure traits on risk of cardiovascular diseases: GWAS and Mendelian randomisation in 100,000 Chinese adults": Supplementary_Figure14.docx

**Supplementary Figure 14. Multivariable Mendelian randomisation of blood pressure with major cardiovascular diseases and subclinical atherosclerosis**. Effects are shown as odds ratios (95% CI) of disease risk per 1 SD higher blood. Loci from two traits were merged and in cases where more than one variant was available in the locus, the variant from the first trait was always included as the instrument when constructing the polygenic score. SBP indicates systolic blood pressure; DBP, diastolic blood pressure; PP, pulse pressure; MAP, mean arterial pressure; SV, single variable MR; MV, multi-variable MR.

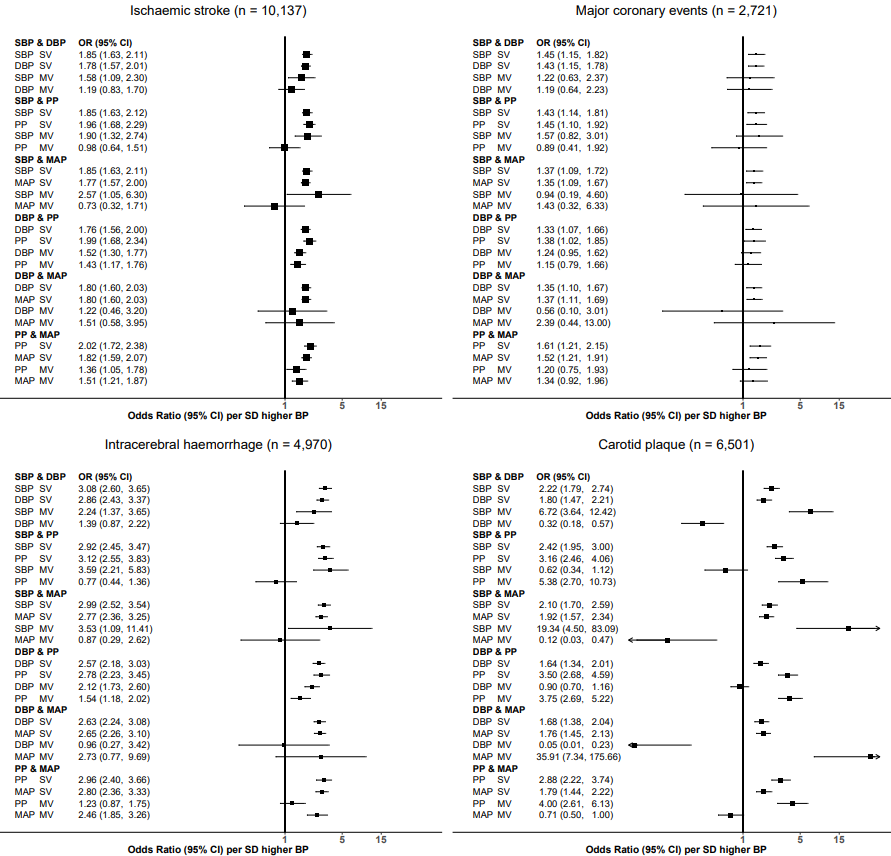
