## Supplementary figures 1-16 for "Causal relevance of different blood pressure traits on risk of cardiovascular diseases: GWAS and Mendelian randomisation in 100,000 Chinese adults": Supplementary_Figure 1.docx

**Supplementary Figure 1. Study flow chart for the China Kadoorie Biobank participants.** Inclusion/exclusion criteria for the GWAS and Mendelian randomisation analyses are summarised.

**All participants in the China Kadoorie Biobank (n=513,214)**

- BMI value missing, n=2
- Any BP trait (i.e., SBP, DBP, PP and MAP) data points > 5 SDs from the mean, n=371

**Participants after removal of outliers and missing data (n=512,841)**

- Any BP trait (i.e., SBP, DBP, PP and MAP) data points > 5 SDs from the mean of residuals, n=411
- Not genotyped, n=411,977

**Genotyped (n=100,453)**

GWAS of BP traits in the whole population

Genetic outliers within each study region (n=6,096)

**Participants after removal of genetic outliers (n=94,357)**

Meta-analysis of BP traits

**Mendelian Randomisation**

Self-reported prior CVD

(n=3,942)

Common controls

(n=72,587)

Ischaemic stroke

(n=10,137)

Intracerebral haemorrhage

(n=4,970)

Major coronary events

(n=2,721)

Carotid plaque

(n=6,501)

**First vascular event**

BP, blood pressure, including systolic, diastolic, pulse and mean arterial blood pressure (SBP, DBP, PP, and MAP); BMI, body mass index; CVD, cardiovascular disease; COPD, chronic obstructive pulmonary disease; CIMT, carotid intima-media thickness.
