## Supplementary figures and images for "Causal relevance of different blood pressure traits on risk of cardiovascular diseases: GWAS and Mendelian randomisation in 100,000 Chinese adults"

### Supplementary_Figure16.docx

**Supplementary Figure 16. Analysis workflow for replication of novel SNPs in BioBank Japan.**

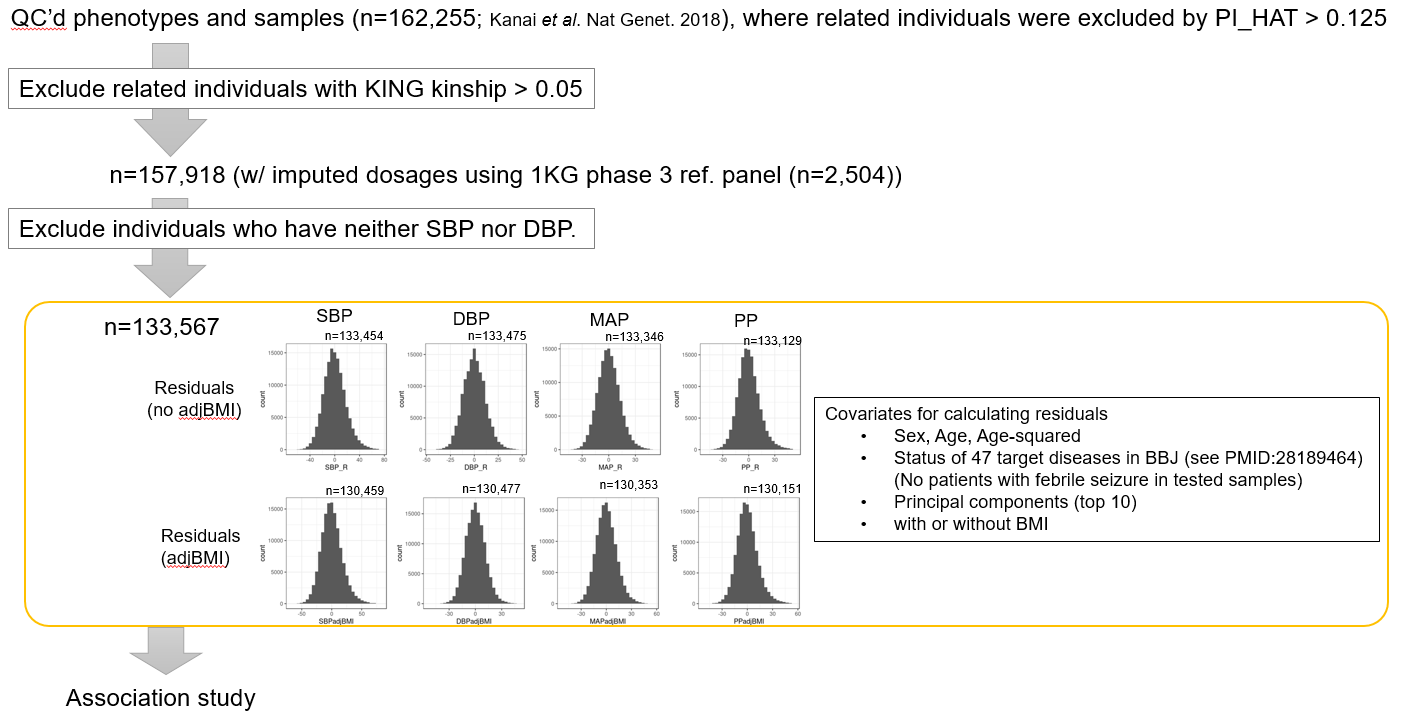
