## Supplementary information for "Causal relevance of different blood pressure traits on risk of cardiovascular diseases: GWAS and Mendelian randomisation in 100,000 Chinese adults"

**International Co-ordinating Centre, Oxford:** Daniel Avery, Derrick Bennett, Ruth Boxall, Sue Burgess, Ka Hung Chan, Yiping Chen, Zhengming Chen, Johnathan Clarke; Robert Clarke, Huaidong Du, Ahmed Edris Mohamed, Zammy Fairhurst-Hunter, Hannah Fry, Simon Gilbert, Mike Hill, Pek Kei Im, Andri Iona, Maria Kakkoura, Christiana Kartsonaki, , Kuang Lin, Mohsen Mazidi, Iona Millwood, Sam Morris, Qunhua Nie, Alfred Pozarickij, Paul Ryder, Saredo Said, Sam Sansome, Dan Schmidt, Paul Sherliker, Rajani Sohoni, Becky Stevens, Iain Turnbull, Robin Walters, Lin Wang, Neil Wright, Ling Yang, Xiaoming Yang, Pang Yao.

**National Co-ordinating Centre, Beijing:** Yu Guo, Xiao Han, Can Hou, Qingmei Xia, Chao Liu, Jun Lv, Pei Pei, Canqing Yu.

**10 Regional Co-ordinating Centres: Guangxi** Provincial CDC: Naying Chen, Duo Liu, Zhenzhu Tang. Liuzhou CDC: Ningyu Chen, Qilian Jiang, Jian Lan, Mingqiang Li, Yun Liu, Fanwen Meng, Jinhuai Meng, Rong Pan, Yulu Qin, Ping Wang, Sisi Wang, Liuping Wei, Liyuan Zhou. **Gansu** Provincial CDC: Caixia Dong, Pengfei Ge, Xiaolan Ren. Maiji CDC: Zhongxiao Li, Enke Mao, Tao Wang, Hui Zhang, Xi Zhang. **Hainan** Provincial CDC: Jinyan Chen, Ximin Hu, Xiaohuan Wang. Meilan CDC: Zhendong Guo, Huimei Li, Yilei Li, Min Weng, Shukuan Wu. **Heilongjiang** Provincial CDC: Shichun Yan, Mingyuan Zou, Xue Zhou. Nangang CDC: Ziyan Guo, Quan Kang, Yanjie Li, Bo Yu, Qinai Xu. **Henan** Provincial CDC: Liang Chang, Lei Fan, Shixian Feng, Ding Zhang, Gang Zhou. Huixian CDC: Yulian Gao, Tianyou He, Pan He, Chen Hu, Huarong Sun, Xukui Zhang. **Hunan** Provincial CDC: Biyun Chen, Zhongxi Fu, Yuelong Huang, Huilin Liu, Qiaohua Xu, Li Yin. Liuyang CDC: Huajun Long, Xin Xu, Hao Zhang, Libo Zhang. **Jiangsu** Provincial CDC: Jian Su, Ran Tao, Ming Wu, Jie Yang, Jinyi Zhou, Yonglin Zhou. Suzhou CDC: Yihe Hu, Yujie Hua, Jianrong Jin Fang Liu, Jingchao Liu, Yan Lu, Liangcai Ma, Aiyu Tang, Jun Zhang. **Qingdao** Qingdao CDC: Liang Cheng, Ranran Du, Ruqin Gao, Feifei Li, Shanpeng Li, Yongmei Liu, Feng Ning, Zengchang Pang, Xiaohui Sun, Xiaocao Tian, Shaojie Wang, Yaoming Zhai, Hua Zhang, Licang CDC: Wei Hou, Silu Lv, Junzheng Wang. **Sichuan** Provincial CDC: Xiaofang Chen, Xianping Wu, Ningmei Zhang, Weiwei Zhou. Pengzhou CDC: Xiaofang Chen, Jianguo Li, Jiaqiu Liu, Guojin Luo, Qiang Sun, Xunfu Zhong. **Zhejiang** Provincial CDC: Weiwei Gong, Ruying Hu, Hao Wang,Meng Wan, Min Yu. Tongxiang CDC: Lingli Chen, Qijun Gu, Dongxia Pan， Chunmei Wang, Kaixu Xie, Xiaoyi Zhang.
